# Measuring the health of people in places: a scoping review of OECD member countries

**DOI:** 10.1101/2021.04.14.21255454

**Authors:** Emily T Murray, Nicola Shelton, Paul Norman, Jenny Head

## Abstract

**Background:** Defining and measuring population health in places is fundamental for local and national planning and conducting within-country and cross-national health comparisons. Yet availability and comparability of place-level health data is unknown.

**Methods:** A scoping review was performed to identify how Organisation for Economic Co-operation and Development (OECD) countries measure overall health for sub-national geographies within each country. The search was conducted across MEDLINE, Scopus and Google Scholar, supplemented by searching all 38 OECD countries statistical agency and public health institute websites.

**Results:** Sixty publications were selected, plus extracted information from 37 of 38 OECD countries statistical agency and/or public health institute websites. Data sources varied by categorisation into mortality (n=7) or morbidity (n=5) health indicators: the former mostly from national statistical agencies and the latter from population-level surveys. Region was the most common geographic scale: eight indicators for 26 countries, two indicators for 24 countries and one indicator for 20 countries. Similar but slightly fewer indicators were available for urban areas (max countries per most frequent indicator = 24), followed by municipality (range of 1-14 countries per indicator). Other geographies, particularly those at smaller granularity, were infrequently available across health indicators and countries.

**Conclusion:** Health indicator data at sub-national geographies are generally only available for a limited number of indicators at large administrative boundaries. Relative uniformity of health indicator question format allows cross-national comparisons. However, wider availability of health indicators at smaller, and non-administrative, geographies is needed to explore the best way to measure population health in local areas.

## INTRODUCTION

It is well known that health varies by geography. Both across countries, within countries, and even within local geographies, people with better and worse health tend to cluster in different locations. These geographic health divides are longstanding and universal.^1^

Whether these spatial health clusters, or what we call here ‘health in a place’, reflect causal processes, or are just an artefact of people with similar health states tending to live in the same places, is currently being debated in the scientific literature.^2^ What is important on a practical level is being able to document and measure these spatial health clusters not just for research purposes, but also for local, national and cross-national planning for health provision, social care, welfare spending and community services; to name a few. Arguments have also been made that the health of people in places as a whole should be viewed as a social and economic asset in its own right.^3^

One of the fundamental exercises in this process is to define which health indicators should be used to measure health in a population. Many scholars and civil servants have given time and thought to this complex issue. At a core level, there are theoretical considerations of what is a healthy or unhealthy population,^4, 5^ dominated by whether definitions should only include normal biological functioning or be expanded to include complete wellbeing.^6^ In practice, theoretical definitions of health indicators tend to give way to which indicators are useful to governments and/or institutions for population health monitoring, policy formation and evaluation.^5, 7^ For example, the World Health Organisation (WHO) defines health as a “state of complete physical, mental and social well-being and not merely the absence of disease or infirmity.”^8^ Yet the official WHO core health indicators contain many disease-specific indicators,^9^ with a new classification system introduced for the ‘measurement’ of health, called The International Classification of Functioning, Disability and Health, endorsed in 2001 by all Member States.^10^ Other methods used by organisations to try to meld these different priorities into a core set of required health indicators have been to develop formal assessment tools,^11^ expert panels^12^ and co-produced health indices.^13-15^ The reality being that many different organisations collect many different population-level health indicators for many different reasons.

When measuring health in a place, an added complication is which geographic boundary, or boundaries, should be used. Here too the theoretical and conceptual struggles meld with the practical. Generally, neighbourhood effects researchers would prefer smaller, and potentially ‘bespoke’ spatial definitions, to reflect that many health-related socio-spatial processes occur at local levels that vary by individual perception and space usage.^16^ Some researchers have recently argued for taking into consideration larger political and economic structures when investigating links between health and place.^17^ The latter would align better with the needs of national and international requirements for public health monitoring, policy development, administrative funding allocation, planning health services and programme evaluation; to name a few.^5^ We must acknowledge however that the mechanisms that influence health of people in places, and collective population health relationships with higher-level social and economic inequalities, are most likely occurring at multiple geographic scales simultaneously.^16^ Whether current health indicator data at sub-national geographies is available to meet the needs of these multiple parties is currently unknown.

## METHODS

The review follows the four-stage approach of Arksey and O’Malley,^18^ detailed below:

### (1) Identifying the research questions

The overall objective of this scoping review was to systematically identify which health indicators are available at sub-national geographies for countries in the OECD. This is in order to answer the following research questions: (1) Which overall health indicators are being used to represent health in a place, (2) what geographic boundary size(s) are being used to represent place when examining population health, (3) does the indicator represent health for all ages in a population and (4) where the health indicator data can be obtained.

### (2) Identifying relevant studies

English-language publications were identified by searching electronic databases: Ovid Medline and Scopus for journal articles and Google Scholar for grey literature (e.g. public health reports), for publication years 2010 to 2020. First, the search strategy for Ovid Medline (Supplementary Table S1) was developed by ETM and modified in discussion with JH, NS and PN. Three concepts of ‘health indicator’, ‘population assessment’ and ‘OECD countries’ were used to only identify studies where health indicators have been used to assess population health at sub-national geographies for the 38 countries currently members of the Organisation for Economic Co-operation and Development.^19^ The OECD search filter was adapted from the Canadian Health Libraries Association.^20^

To evaluate the search strategy, a random 100 publications identified through the Ovid MEDLINE database were both independently screened by ETM and JH on the basis of title and abstract. Reasons for exclusion included the following: (1) the study was not conducted in an OECD country, (2) there was no overall health indicator available (e.g. only a component of health, a health behaviour or syndromic surveillance), (3) no population-level assessment of the health indicator at sub-national geographies (e.g. sub-group assessment only, only one local area) and/or (4) no data assessed (e.g. editorial).

The agreed search strategy was then applied by ETM to the remaining Ovid Medline search results, the Scopus search (see Supplementary Table 2) and to the Google Scholar search (see Supplementary Table S3). Endnote X10 was used to import and manage all publications. For Google Scholar, the search interface necessitated conducting each combination of key words within the ‘health indicator’ and ‘population assessment’ concepts separately and only importing records to Endnote that had been screened initially by title only.

### (3) Study selection

Next, all included articles by title and abstract were assessed for further eligibility by full-text assessment. Due to the difficulty of assessing abstracts for whether health indicators are available at a sub-national geographic level, the geographic criteria were only applied at this stage. Identified study websites were also visited to check for updated information.

In addition to a traditional literature search, we conducted key word searches using the internet search engine Google to identify English-language statistical institutes, national public health institutes and health ministry websites of the 38 members of the OECD countries. Initially, each website was assessed for availability of overall health indicators, followed by whether health indicators were available at sub-national geographic levels.

### (4) Data Extraction

Data from included studies and data sources were extracted in a uniform manner. We extracted the following information: country, data source, data collected year(s), number of indicators, how indicators were measured, the geographic levels at which indicators were available and reference information (i.e. citation information for publications and hypertext links for statistical agency data). Notes were also kept on whether data was fed into other data sources (e.g. national health surveys that are a part of the European Health Examination Survey). Data were extracted by one reviewer (ETM) and a second reviewer (JH) performed an independent data extraction for a randomly chosen 10% of publications (n=6). Inconsistent results were discussed, and the extraction modified accordingly.

## RESULTS

For the initial literature review, we identified 60 publications (Figure 1). At the title/abstract screening stage (n=1,157 non-duplicates), the most common reasons for exclusion was the health indicator(s) did not cover overall health (n=459) (e.g. a health component indicator, a health behaviour indicator, etc) or the overall health indicator was not available at a population level (n=303)(e.g. assessed sub-population groups only, only assessed one locality, etc), followed by the publication being an editorial piece only (n=95), data not available in an OECD country (n=71) and no full text available to review (n=18)(e.g. conference presentation abstract only). For the 210 full texts reviewed, a third (56 out of 150) were excluded for the health indicators not having been available at a sub-national geography. The remaining exclusions were distributed similarly as the title/abstract screening stage.

**Figure 1.**
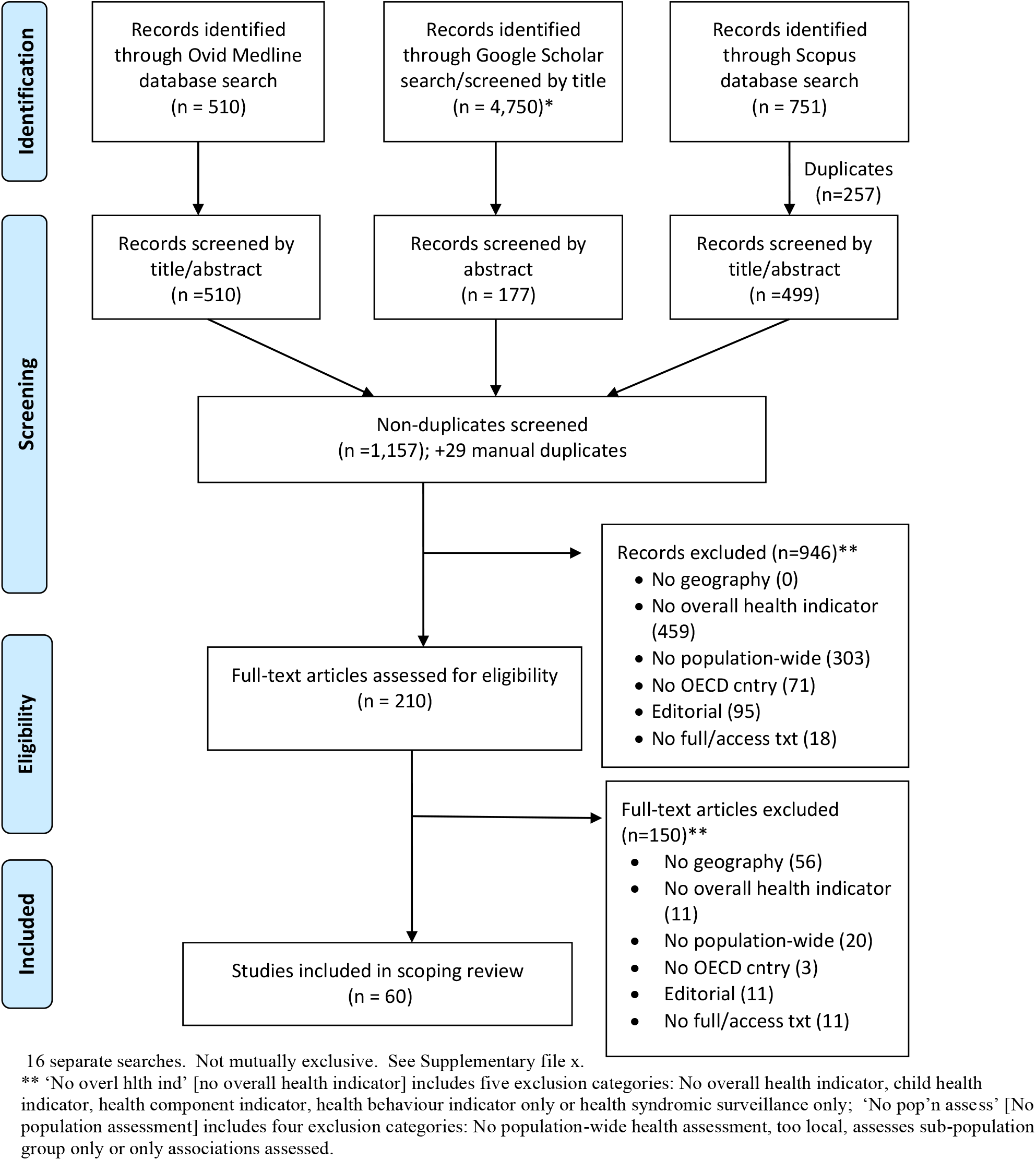
Flow Diagram for selection of scoping review articles (2010-2020) 16 separate searches. Not mutually exclusive. See Supplementary file x. ** ‘No overl hlth ind’ [no overall health indicator] includes five exclusion categories: No overall health indicator, child health indicator, health component indicator, health behaviour indicator only or health syndromic surveillance only; ‘No pop’n assess’ [No population assessment] includes four exclusion categories: No population-wide health assessment, too local, assesses sub-population group only or only associations assessed.

For almost all OECD countries, with the exception of Chile, additional information on overall health indicator data was available either on the country’s statistical agency or public health institute English-language section(s) of their website(s). For studies that had been identified during the literature review, study websites were assessed for further information. Specific information on where health indicator data were identified for each country are located in Supplementary Table 4. For all countries investigated, a comprehensive understanding of available health indicators could not be obtained by academic search engines alone. Even when a specific data source was used in a study (e.g. mortality records), additional information could usually be found on the specific country or study website.

### Health Indicators by Data Source

Mortality and morbidity indicators were generally obtained from different sources, so will be presented separately.

Table 1 summarises which mortality indicators were available at a population level below country-level, including the health indicator data source, year(s) of data collection and geographic data boundar(ies) the mortality indicators were available. For the 38 OECD countries, all mortality indicators were available from governmental statistical or public health institutes. The timeframe and years of data collection were highly variable by country. Six organisations or studies compiled all-cause mortality data for sub-national geographic boundaries across multiple countries: OECD.stat (38 countries) for Territorial Levels 2 and 3 boundaries (OECD sub-national administrative classification tiers: TL2 ‘394 larger regions’ and TL3 ‘2,258 smaller regions’),^21^ Eurostat Weekly deaths (28 countries)^22^ and EURO-HEALTHY (28 countries)^12, 13, 23, 24^ for NUTS 2 (European Union ‘Nomenclature of territorial units for statistics,’ 281 regions),^25^ EURO-URHIS2 (14 countries)^26-28^ for project-specific urban areas, and WHO European Healthy cities (all European countries).^29^ EURO-HEALTHY also compiles cause-specific mortality, life expectancy at birth and preventable mortality for 28 European regions,^12, 13, 23, 24^ while EURO-URHIS2 only additionally calculates cause-specific mortality for their specified urban areas.^26-28^ The EuroMOMO study releases regional-level excess mortality data for 24 European countries, calculated from each country’s weekly official national mortality statistics.^30, 31^

For morbidity indicators, 37 OECD countries’ data (excluding Israel) were available in English for sub-national geographies, but varied widely by data source, timescale of data availability, age range of the sample, morbidity indicator and geographic scale. Therefore, Table 2 summarizes sub-national geographic data availability for each OECD country by data source category. Again, a number of studies – EURO-HEALTHY, EU-SILC, EHIS and EURO-URHIS - have morbidity indicator data available for multiple European countries for sub-national geographies; generally for regions, municipalities and/or urban areas.^12, 13, 23, 24, 26-28, 32, 33^ For non-European OECD countries, and additional data collection by European countries, morbidity indicator data is available by other Health Interview Surveys (i.e. not a part of EURO-HEALTHY, EU-SILC, EHIS or EURO-URHIS), Health Examination Surveys, Other (not necessarily health) surveys and/or Censuses (see Supplementary Table 4). Within these additional surveys, health indicator availability at sub-national geographies varied considerably. See Supplementary Table 5 for availability of each morbidity indicator, and associated sub-national geographic scale, for each country’s specific data source.

**Table 1.**
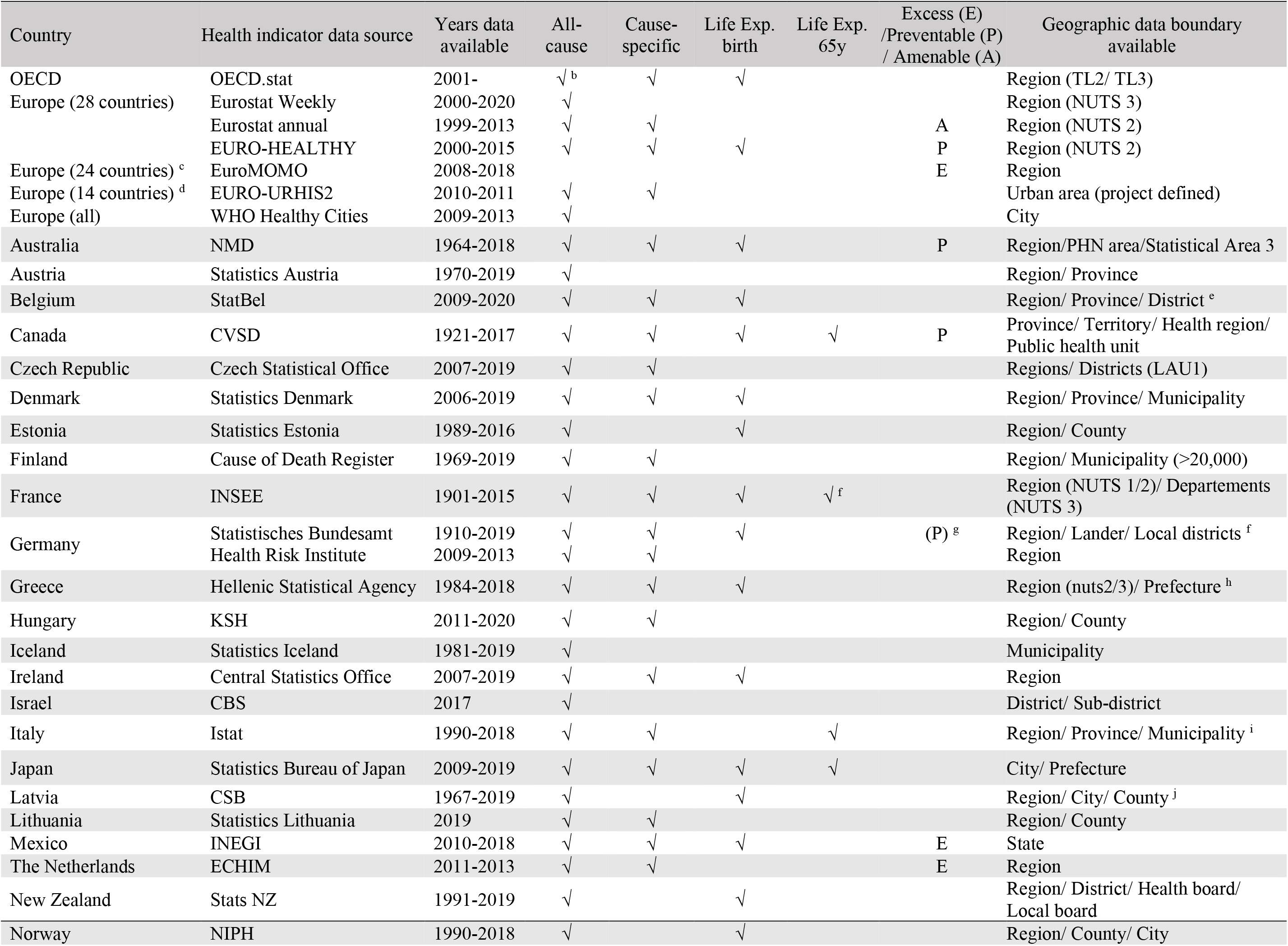
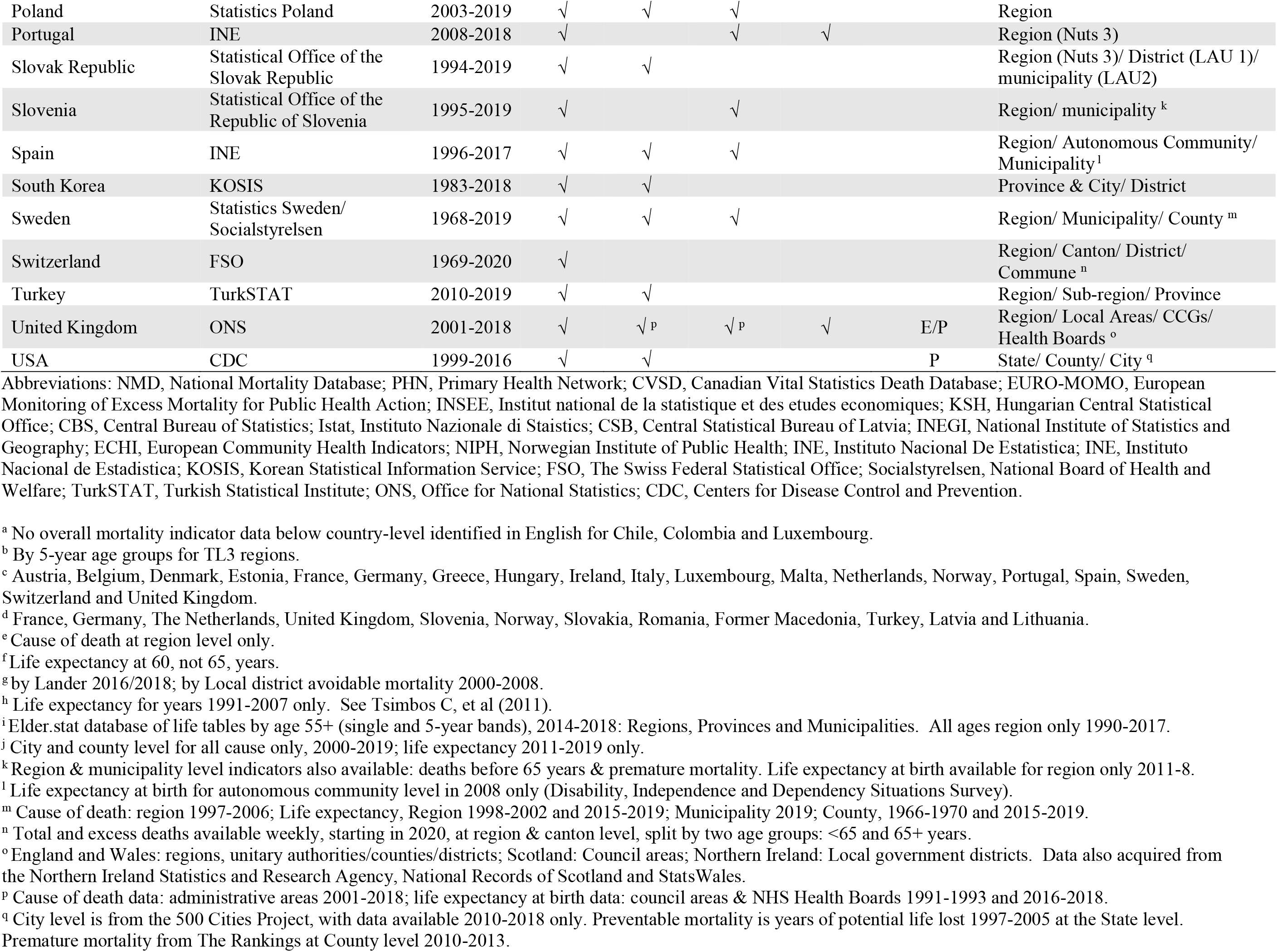
Health indicators available in OECD countries a by sub-country geographies: mortality.

**Table 2.** Data source(s) for morbidity indicators by country and geographic boundary.

|  |  |  |  |  |  |  |  |  |  |  |
| --- | --- | --- | --- | --- | --- | --- | --- | --- | --- | --- |
|  | EURO-HEALTHY | EU-SILC | EHIS | EHIS2 | EURO-URHIS | EURO-URHIS2 | Other HIS | Health Exam<br>Survey | Other Survey(s) | Census |
| Australia |  |  |  |  |  |  | S/ T |  |  | Many |
| Austria | R2 | R1 | R2 | R2 | UA |  |  |  |  |  |
| Belgium | R2/ Mu | R1 | R2 | R2 | UA |  | R/ P |  |  |  |
| Canada |  |  |  |  |  |  | P/ T/ MA |  | P/ T/ MA |  |
| Chile |  |  |  |  |  |  |  | R |  |  |
| Colombia |  |  |  |  |  |  |  | R | R |  |
| Czech Republic | R2/ Mu | R1 | R2 | R2 | UA |  |  |  |  |  |
| Denmark | R2 | R1 |  | R2 | UA |  |  |  |  |  |
| Estonia | R2 | R1 | R2 | R2 | UA |  |  |  |  | R |
| Finland | R2 | R1 |  | R2 | UA |  |  | R/ Mu |  |  |
| France | R2/ Mu | R1 | R2 | R2 | UA | UA |  |  |  |  |
| Germany | R2/ Mu |  |  | R2 | UA | UA | R/ Mu | R | R |  |
| Greece | R2/ Mu | R1 |  | R2 | UA |  |  |  |  |  |
| Hungary | R2 | R1 |  | R2 | UA |  |  |  |  | R/ Co |
| Iceland | R2 | R1 |  | R2 | UA |  |  |  |  |  |
| Ireland | R2 | R1 |  | R2 | UA |  |  |  |  | R/ Co/ City |
| Israel |  |  |  |  |  |  |  |  |  |  |
| Italy | R2/ Mu | R1 |  | R2 | UA |  |  |  | Mu |  |
| Japan |  |  |  |  |  |  |  |  | Di/Prefecture |  |
| Korea |  |  |  |  |  |  | Co/ Di/ City |  |  |  |
| Latvia | R2 | R1 | R2 | R2 | UA | UA |  |  |  |  |
| Lithuania | R2 | R1 |  | R2 | UA | UA |  |  | R/ Co |  |
| Luxembourg | R2 | R1 |  | R2 |  |  |  |  |  |  |
| Mexico |  |  |  |  |  |  |  | Admin T |  |  |
| Netherlands | R2 | R1 |  | R2 | UA | UA |  |  | Postcode |  |
| New Zealand |  |  |  |  |  |  | R/ Di/ PHU |  |  | R |
| Norway | R2 | R1 |  | R2 | UA | UA |  |  |  |  |
| Poland | R2 | R1 |  | R2 | UA |  |  |  |  |  |
| Portugal | R2/ Mu | R1 |  | R2 |  |  |  | R/ UA |  |  |
| Slovak Republic | R2 | R1 |  | R2 | UA | UA |  |  | R2 |  |
| Slovenia | R2 | R1 | R2 | R2 | UA | UA |  |  |  |  |
| Spain | R2/ Mu | R1 |  | R2 | UA |  | AC/ Mu |  |  |  |
| Sweden | R2/ Mu | R1 |  | R2 | UA |  |  |  |  |  |
| Switzerland | R2 | R1 | R2 |  |  |  |  |  |  |  |
| Turkey |  | R1 | R2 |  | UA | UA |  |  |  |  |
| United Kingdom | R2/ Mu | R1 |  | R2 | UA | UA | R/ LA/ HB | R | R | LA/ OA/ DZ |
| United States |  |  |  |  |  |  | Many |  | State/Co/Ma/Mi | Many |

Abbreviations: AU, Autonomous Community; Co, County; Di, District; DZ, Data Zone; HB, Health Board; HIS, Health Interview Survey; LA, Local Authority; MA, Metropolitan Area; Mu, Municipality; P, Province; PHU, Public Health Unit; R, Region not NUTS or unspecified; R1, Region NUTS 1; R2, Region NUTS2; OA, Output Area; T, Territory; UA, Urban Area.

### Health Indicators by Geographic level frequency

Table 3 summarizes the frequency that health indicators were identified at sub-national geographies across OECD countries. For example, only one country was identified to have NUTS1 (major socio-economic regions)^25^ geographic level data on all-cause mortality, cause-specific mortality, life expectancy at birth, life expectancy at age 65 years and disability. In contrast, 26 OECD countries had data on self-rated health, long-standing illness and activity limitation at the same aggregate geographic level. Overall, ‘region (NUTS 2)’^25^ was the most common geographic boundary where health indicator data was available. Three of the eight identified health indicators were available for all 38 OECD countries, six of the twelve were available for 26 countries, two indicators (cause-specific mortality and healthy life expectancy) were available for 24 countries and one indicator (excess mortality) was available for 20 countries. The second most frequent geographic level was ‘urban area’, with data from 24 countries for five common health indicators (23 for long-standing illness). Health indicator data was also available frequently at ‘municipality’ level, with nine of the indicators available at this geographic level for a low of seven, and high of 14, OECD countries. Health indicator data below municipality, or equivalent geographic size, and at any geography for life expectancy at age 65 years, was sparse. For a listing of sub-national geographic availability for specific OECD countries, see Supplementary Table 6.

**Table 3.** Summary of overall health indicator availability by OECD country and sub-country geography.

|  |  |  |  |  |  |  |  |  |  |  |  |  |
| --- | --- | --- | --- | --- | --- | --- | --- | --- | --- | --- | --- | --- |
|  | Mortality Indicators |  |  |  |  |  |  | Morbidity Indicators |  |  |  |  |
|  | All-cause | Cause-specific | Life Exp birth | Life Exp 65y | Preventable | Excess | Amenable | Self-rated health | Long-standing illness | Activity limitation | Disability | Healthy Life Expectancy |
| **Large geographies:** |  |  |  |  |  |  |  |  |  |  |  |  |
| Region (NUTS1) | 1 | 1 | 1 | 1 | 0 | 0 | 0 | 26 | 26 | 26 | 1 | 0 |
| Region (NUTS2/TL2) | 38 | 38 | 38 | 2 | 26 | 20 | 26 | 26 | 26 | 26 | 26 | 24 |
| Autonomous Community (Spain) | 0 | 1 | 0 | 0 | 0 | 0 | 0 | 0 | 0 | 0 | 0 | 0 |
| Region (Unspecified) | 2 | 1 | 1 | 1 | 1 | 0 | 0 | 2 | 1 | 2 | 0 | 0 |
| Public Health Region | 0 | 1 | 1 | 1 | 1 | 0 | 0 | 0 | 0 | 0 | 0 | 0 |
| Province | 7 | 3 | 1 | 1 | 1 | 0 | 0 | 1 | 0 | 1 | 1 | 1 |
| Territory | 1 | 1 | 1 | 1 | 1 | 0 | 0 | 3 | 2 | 1 | 2 | 1 |
| State | 2 | 2 | 1 | 0 | 0 | 1 | 0 | 2 | 1 | 1 | 2 | 0 |
| **Medium Geographies:** |  |  |  |  |  |  |  |  |  |  |  |  |
| Region (NUTS3/TL3) | 38 | 3 | 38 | 2 | 0 | 0 | 0 | 0 | 0 | 0 | 0 | 0 |
| Sub-Region | 0 | 1 | 0 | 0 | 0 | 0 | 0 | 0 | 0 | 0 | 0 | 0 |
| Prefecture | 2 | 1 | 1 | 1 | 0 | 0 | 0 | 0 | 0 | 0 | 0 | 0 |
| County | 7 | 4 | 4 | 0 | 0 | 0 | 0 | 3 | 1 | 2 | 3 | 0 |
| Local Authority (UK) | 1 | 1 | 1 | 1 | 1 | 1 | 0 | 1 | 1 | 1 | 1 | 1 |
| Metropolitan Area | 0 | 0 | 0 | 0 | 0 | 0 | 0 | 2 | 0 | 2 | 1 | 0 |
| Urban Area | 24 | 24 | 0 | 0 | 0 | 0 | 0 | 24 | 23 | 24 | 0 | 0 |
| City | 5 | 3 | 3 | 1 | 0 | 0 | 0 | 2 | 0 | 1 | 1 | 0 |
| District | 6 | 4 | 2 | 0 | 1 | 0 | 0 | 4 | 1 | 1 | 1 | 0 |
| Municipality | 14 | 13 | 11 | 1 | 9 | 1 | 10 | 12 | 12 | 7 | 11 | 2 |
| Commune | 0 | 1 | 0 | 0 | 0 | 0 | 0 | 0 | 0 | 0 | 0 | 0 |
| **Small geographies:** |  |  |  |  |  |  |  |  |  |  |  |  |
| UK Output Area/Data Zone (UK) | 1 | 0 | 0 | 0 | 1 | 0 | 0 | 1 | 1 | 0 | 1 | 0 |
| Public Health Unit | 1 | 1 | 1 | 1 | 1 | 0 | 0 | 0 | 0 | 0 | 0 | 0 |
| Clinical Commissioning Group (UK) | 1 | 1 | 1 | 1 | 1 | 1 | 0 | 0 | 0 | 0 | 0 | 0 |
| Health Board (NZ/UK) | 1 | 0 | 1 | 1 | 1 | 1 | 0 | 0 | 0 | 0 | 0 | 0 |
| Census Tract (US) | 0 | 0 | 0 | 0 | 0 | 0 | 0 | 1 | 0 | 0 | 1 | 0 |
| Census Block Group (US) | 0 | 0 | 0 | 0 | 0 | 0 | 0 | 1 | 0 | 0 | 1 | 0 |
| Postcode | 0 | 0 | 0 | 0 | 0 | 0 | 0 | 1 | 1 | 0 | 0 | 0 |
| Census Block (US) | 0 | 0 | 0 | 0 | 0 | 0 | 0 | 1 | 0 | 0 | 1 | 0 |

Abbreviations: AC, Autonomous Community; Co, County; Di, District; DZ, Data Zone; HB, Health Board; HIS, Health Interview Survey; LA, Local Authority; Ma, Metropolitan Area; Mu, Municipality; P, Province; PHU, Public Health Unit; R, Region not NUTS or unspecified; R1, Region NUTS 1; R2, Region NUTS2; OA, Output Area; St, State; T, Territory; UA, Urban Area.

### Health Indicators by Population Age

Of the 12 health indicators identified in this review, only one, life expectancy at age 65, addressed a specific age range. This indicator was only available at sub-national geographies for six of the OECD countries: Canada (Province/Territory, Public Health Region and Public Health Unit), France (Region: NUTS 1/2/3), Italy (Region, Prince and Municipality), Japan (City and Prefecture), Portugal (NUTS 3) and the UK (Region NUTS 2, Local Authority, Clinical Commissioning Group and Health Board) (see Supplemental Table 6).

Except for life expectancy at age 65, sub-national mortality data generally represented the entire age range of the population. For sub-national morbidity indicators, the age range varied by, and within, the data source. Health indicators from census data covered the entire population. Some surveys would cover the entire age range of the population, while others only cover an ‘adult’ population, but the age where adulthood began mostly ranged from age 15 to 25 years. Exceptions were the Canadian Community Health Survey (CCHS), age range 12+, and the Spanish Instituto Nacional de Estadistica (INE), with an age range 9+. We only identified one study with sub-national health indicator data for only older people (age 60+): Columbia’s Survey on Health, Well-being and SAlud (SABE) (see Supplementary Table 5).

## DISCUSSION

In this comprehensive scoping review of academic journal articles, grey literature and government statistical & public health websites, health indicator availability for sub-national geographies were limited in both number, data source and geographic scale. Across the 38 OECD countries, only twelve overall health indicators were available at a population level for sub-national geographies, seven mortality and five morbidity. Region, or equivalent large subnational entities, was the predominant geographic level for both mortality and morbidity indicators. Health indicator availability at smaller geographies was sparse, and varied considerably by geographic definition, health indicator, age range of population and years available. In all cases, geographic boundaries used only administrative definitions.

The finding that only a dozen health indicators were available at any sub-national geographies is most likely a result of several cross-national initiatives to harmonize health indicators at larger geographies. Historically, this included the World Health Organization’s (WHO) framework for recorded causes of death and Health for All Programme, plus health indicator data collections by the OECD and Eurostat. The European Union has conducted a series of health indicator harmonisation projects, starting with the Amsterdam Treaty in 1993 and continuing through jointly agreeing to a shortlist of indicators in the mid-2000s and the Joint Action for ECHIM in 2009.^34^ In 2017, experts nominated by EU Member States agreed a set of 40 health indicators for a Joint Monitoring Framework (JMF), which would be used to measure achievement of the Sustainable Development Goals (SDGs), Health 2020 and the Global Action Plan for the Prevention and Control of Non-communicable Diseases (NCDs).^35^ Of these, four reflect overall health: life expectancy at birth, life expectancy at 65, healthy life expectancy and general mortality.

Why regions, specifically the NUTS 2 definition, is the most frequent geographic boundary available for health indicators, is almost surely due to the ISARE (Health Indicators in the European Regions) projects, who led the collection and harmonisation of health indicator data at NUTS 2 regional levels.^36^ In addition, in the EU the NUTS 2 level designation is used by the EU Commission to allocate funds.^11, 23^ The importance of these regions as political and administrative units,^11^ particularly for healthcare funding and planning, would similarly explain why health indicators are routinely collected at similar large sub-national geographies in other non-European countries (e.g. States in USA, Provinces/Territories in Canada, etc).^37^

Why health indicator data is not more frequently available at smaller, and/or non-administrative geographic scales is unclear. All-cause mortality is available at ‘a’ local geographic level for most OECD countries, but the population size and/or spatial size of these local areas varies widely. Indicators of cause-specific mortality, life expectancy at birth and particularly life expectancy at age 65 at smaller geographies could be mostly calculated from the same data sources of all-cause mortality,^38^ so it is unclear why they are not. Speculative reasons include issues of data access, staff capacity, suppression to protect confidentiality due to small cell counts for age by sex by cause of death, prioritization and/or perceived usefulness of data. For morbidity indicators, the most likely explanation is a lack of many national surveys to sample sufficient participants at a local level to produce reliable local estimates.^33^ Potential solutions include using only highly-dense geographic units,^33^ increasing sample sizes in national surveys to be locally representative (such as the Korean Community Health Survey Profiles^39^), introduce more health questions into national censuses and/or develop new potential big data technologies, such as electronic health records.^40^

The lack of sub-national health indicators or data sources for specific age groups, particularly older people, is concerning. The lack of data sources specific to older populations with sub-national health indicator data appears to be attributable to most ageing studies sampled to be only nationally, and not locally, representative.^41^ Equally, we speculate that national surveys with sub-national health indicator data may not have large enough sample sizes in specific age ranges that could be representative and/or released without disclosing personal information of participants.

The main strength of this paper is for the first time creating a summary of overall health indicator data at sub-national geography for over three-dozen countries. The use of varied publication types makes us confident that the review is comprehensive. In particular, assessing research study and statistical agency websites was a valuable activity. Relying on published journal articles alone would have created an incomplete assessment. The largest limitations were to restrict sources to English language and OECD countries. During the search it was apparent that some OECD countries do produce additional publications in a language other than English. All non-native English-speaking countries also provided English-language versions of their websites. It is however unknown if additional information on health indicator data can be found on the non-English websites. We could have contacted representatives from each country, but a decision was made to have the review reflect information and data that was publicly available for comparison purposes. For the health indicator assessment, we chose to focus on overall, rather than component or behavioural health indicators, as well as adult rather than childhood health indicators. This reflected the decision to not initially exclude publications on geographic criteria, which meant that with each broadening of the concept of health, and increase in age range, the initial search results were excessive. A similar size scoping review could separately be done on each concept of health components, behavioural risk factors, well-being measures and childhood health indicators.

In conclusion, our scoping review has shown that measuring overall health in OECD sub-national populations is restricted mainly to a dozen indicators at large geographies. In one sense, this is positive and reflects decades of health monitoring cooperation and harmonisation by EU countries. On the downside, publicly available data on the health of local populations is sparse, comes from limited data sources, only reflects administrative geographic boundaries and is not comparable across countries. We recommend that health monitoring studies be altered and/or new technologies be designed, to allow increased public health monitoring of health at the local level.

## Data Availability

All data referred to in the manuscript is publicly available. Links are provided in supplementary materials.

