## Supplemental tables 1-6 for "Measuring the health of people in places: a scoping review of OECD member countries"

| | | |
|---|---|---|
| **Supplementary Table 1. Scoping Review searching strategy for Ovid Medline database.** | | |
| **CONCEPT: HEALTH INDICATORS**<br>( TITLE-ABS-KEY ( "Health Status Indicators" ) OR TITLE-ABS ( "health indicator*" ) ) OR TITLE-ABS ( "health index" ) OR TITLE-ABS ( "health indices" ) OR TITLE-ABS ( "morbidity indicator*" ) OR TITLE-ABS ( "mortality indicator*" ) | | |
| N=32,882 | | |
| **CONCEPT: POPULATION ASSESSMENT**<br>TITLE-ABS-KEY ( "population surveillance" ) OR KEY ( "*Public health" ) OR KEY ( "Population health" ) OR TITLE-ABS ( "community health" ) OR TITLE-ABS ( "surveillance system" ) OR TITLE-ABS ( "public health monitoring" ) OR TITLE-ABS ( "health interview survey*" ) OR TITLE-ABS ( "health examination survey*" ) OR TITLE-ABS ( "health monitoring" )<br><br>N=474,510 | | |
| 3 | 1 and 2 | 3925 |
| **CONCEPT: OECD COUNTRIES**<br>((TITLE-ABS(Canada* or Canadi* or Alberta* or Calgary* or Edmonton* or "British Columbia*" or Vancouver* or Victoria* or Manitoba* or Winnipeg* or "New Brunswick*" or Fredericton* or Moncton* or Newfoundland* or "New Foundland*" or Labrador* or "St John*" or "Saint John*" or "Northwest Territor*" or Yellowknife* or "Nova Scotia*" or Halifax* or Dalhousie* or Nunavut* or Igaluit* or Ontario* or Ontarian* or Toronto* or Ottawa* or Hamilton or Queen's or McMaster* or Kingston* or Sudbury* or "Prince Edward Island*" or Charlottetown* or Quebec* or Montreal* or McGill* or Laval* or Sherbrooke* or Nunavik* or Kuujjuaq* or Inukjuak* or Puvirnituq* or Saskatchewan* or Saskatoon* or Yukon* or Whitehorse* or America* or USA* or "United States*" or "New York*" or Chicago* or Boston* or "San Francisco*" or "Los Angeles*" or "New Orleans*" or Philadelphia* or Chile* or Mexico* or Europe* or Austria* or Belgium* or "Czech Republic*" or France* or Paris* or Germany* or Berlin* or "Great Britain*" or Ireland* or England* or London* or Scotland* or Wales* or "United Kingdom*" or Greece* or Athens* or Hungary* or Italy* or Rome* or Netherlands* or Luxembourg* or Poland* or Portugal* or Scandinav* or Denmark* or Estonia* or Finland* or Iceland* or Norway* or Sweden* or "Slovak Republic*" or Slovenia* or Spain* or Switzerland* or Turkey* or Israel* or Australia* or "New Zealand*" or Japan* or Korea*)) OR (KEY("Organisation for Economic Co-Operation and Development")) OR (TITLE-ABS-KEY("Organisation for Economic Co-Operation and Development"))<br><br>4 N=14,418,318<br><br>5 3 AND 4 = 1,727 | | |

6 LIMIT ‘Human’ = 1,434

7 LIMIT ‘English’ = 1,306

8 LIMIT ‘publication year 2010-2020) = 751

| Supplementary Table 3. Scoping Review searching strategy for Scopus database. | | |
|---|---|---|
| **CONCEPT: HEALTH INDICATORS**<br>( TITLE-ABS-KEY ( "Health Status Indicators" ) OR TITLE-ABS ( "health indicator*" ) ) OR TITLE-ABS ( "health index" ) OR TITLE-ABS ( "health indices" ) OR TITLE-ABS ( "morbidity indicator*" ) OR TITLE-ABS ( "mortality indicator*" ) | | |
| N=32,882 | | |
| **CONCEPT: POPULATION ASSESSMENT**<br>TITLE-ABS-KEY ( "population surveillance" ) OR KEY ( "*Public health" ) OR KEY ( "Population health" ) OR TITLE-ABS ( "community health" ) OR TITLE-ABS ( "surveillance system" ) OR TITLE-ABS ( "public health monitoring" ) OR TITLE-ABS ( "health interview survey*" ) OR TITLE-ABS ( "health examination survey*" ) OR TITLE-ABS ( "health monitoring" )<br><br>N=474,510 | | |
| 3 | 1 and 2 | 3925 |
| **CONCEPT: OECD COUNTRIES**<br>((TITLE-ABS(Canada* or Canadi* or Alberta* or Calgary* or Edmonton* or "British Columbia*" or Vancouver* or Victoria* or Manitoba* or Winnipeg* or "New Brunswick*" or Fredericton* or Moncton* or Newfoundland* or "New Foundland*" or Labrador* or "St John*" or "Saint John*" or "Northwest Territor*" or Yellowknife* or "Nova Scotia*" or Halifax* or Dalhousie* or Nunavut* or Igaluit* or Ontario* or Ontarian* or Toronto* or Ottawa* or Hamilton or Queen's or McMaster* or Kingston* or Sudbury* or "Prince Edward Island*" or Charlottetown* or Quebec* or Montreal* or McGill* or Laval* or Sherbrooke* or Nunavik* or Kuujjuaq* or Inukjuak* or Puvirnituq* or Saskatchewan* or Saskatoon* or Yukon* or Whitehorse* or America* or USA* or "United States*" or "New York*" or Chicago* or Boston* or "San Francisco*" or "Los Angeles*" or "New Orleans*" or Philadelphia* or Chile* or Mexico* or Europe* or Austria* or Belgium* or "Czech Republic*" or France* or Paris* or Germany* or Berlin* or "Great Britain*" or Ireland* or England* or London* or Scotland* or Wales* or "United Kingdom*" or Greece* or Athens* or Hungary* or Italy* or Rome* or Netherlands* or Luxembourg* or Poland* or Portugal* or Scandinav* or Denmark* or Estonia* or Finland* or Iceland* or Norway* or Sweden* or "Slovak Republic*" or Slovenia* or Spain* or Switzerland* or Turkey* or Israel* or Australia* or "New Zealand*" or Japan* or Korea*)) OR (KEY("Organisation for Economic Co-Operation and Development")) OR (TITLE-ABS-KEY("Organisation for Economic Co-Operation and Development"))<br><br>4 N=14,418,318<br><br>5 3 AND 4 = 1,727 | | |

6 LIMIT ‘Human’ = 1,434

7 LIMIT ‘English’ = 1,306

8 LIMIT ‘publication year 2010-2020) = 751

**Supplementary Table 3. Scoping Review searching strategy for Google Scholar database.**

1. "public health" "health indicators" "health monitoring" England OR Wales OR Scotland OR "Northern Ireland" OR “United Kingdom” OR OECD OR Europe OR Canada OR USA OR "United States" OR Austria OR Belgium OR "Czech Republic" OR France OR Germany OR "Great Britain" OR Ireland OR Greece OR Hungary OR Italy OR Netherlands OR Luxemberg OR Poland OR Portugal OR Denmark OR Estonia OR Finland OR Iceland OR Norway OR Sweden OR “Slovak Republic” OR Slovenia OR Spain OR Switzerland OR Turkey OR Israel OR Australia OR “New Zealand” OR Japan OR Korea OR “Organisation for Economic Co-Operation and Development” (n=100/2,240)

2. "population health" AND "health indicators" AND "health monitoring" England OR Wales OR Scotland OR "Northern Ireland" OR “United Kingdom” OR OECD OR Europe OR Canada OR USA OR "United States" OR Austria OR Belgium OR "Czech Republic" OR France OR Germany OR "Great Britain" OR Ireland OR Greece OR Hungary OR Italy OR Netherlands OR Luxemberg OR Poland OR Portugal OR Denmark OR Estonia OR Finland OR Iceland OR Norway OR Sweden OR “Slovak Republic” OR Slovenia OR Spain OR Switzerland OR Turkey OR Israel OR Australia OR “New Zealand” OR Japan OR Korea OR “Organisation for Economic Co-Operation and Development” (n=+41/935)

3. "public health" "mortality indicators" "health monitoring" England OR Wales OR Scotland OR "Northern Ireland" OR “United Kingdom” OR OECD OR Europe OR Canada OR USA OR "United States" OR Austria OR Belgium OR "Czech Republic" OR France OR Germany OR "Great Britain" OR Ireland OR Greece OR Hungary OR Italy OR Netherlands OR Luxemberg OR Poland OR Portugal OR Denmark OR Estonia OR Finland OR Iceland OR Norway OR Sweden OR “Slovak Republic” OR Slovenia OR Spain OR Switzerland OR Turkey OR Israel OR Australia OR “New Zealand” OR Japan OR Korea OR “Organisation for Economic Co-Operation and Development” (n=+2/110)

4. "population health" "mortality indicators" "health monitoring" England OR Wales OR Scotland OR "Northern Ireland" OR “United Kingdom” OR OECD OR Europe OR Canada OR USA OR "United States" OR Austria OR Belgium OR "Czech Republic" OR France OR Germany OR "Great Britain" OR Ireland OR Greece OR Hungary OR Italy OR Netherlands OR Luxemberg OR Poland OR Portugal OR Denmark OR Estonia OR Finland OR Iceland OR Norway OR Sweden OR “Slovak Republic” OR Slovenia OR Spain OR Switzerland OR Turkey OR Israel OR Australia OR “New Zealand” OR Japan OR Korea OR “Organisation for Economic Co-Operation and Development” (n=+0/59)

5. "public health" "morbidity indicators" "health monitoring" England OR Wales OR Scotland OR "Northern Ireland" OR “United Kingdom” OR OECD OR Europe OR Canada OR USA OR "United States" OR Austria OR Belgium OR "Czech Republic" OR France OR Germany OR "Great Britain" OR Ireland OR Greece OR Hungary OR Italy OR Netherlands OR Luxemberg OR Poland OR Portugal OR Denmark OR Estonia OR Finland OR Iceland OR Norway OR Sweden OR “Slovak Republic” OR Slovenia OR Spain OR Switzerland OR Turkey OR Israel OR Australia OR “New Zealand” OR Japan OR Korea OR “Organisation for Economic Co-Operation and Development” (n=+4/37)

6. "population health" "morbidity indicators" "health monitoring" England OR Wales OR Scotland OR "Northern Ireland" OR “United Kingdom” OR OECD OR Europe OR Canada OR USA OR "United States" OR Austria OR Belgium OR "Czech Republic" OR France OR Germany OR "Great Britain" OR Ireland OR Greece OR Hungary OR Italy OR Netherlands OR Luxemberg OR Poland OR Portugal OR Denmark OR Estonia OR Finland OR Iceland OR Norway OR Sweden OR “Slovak Republic” OR Slovenia OR Spain OR Switzerland OR

Turkey OR Israel OR Australia OR “New Zealand” OR Japan OR Korea OR “Organisation for Economic Co-Operation and Development” (n=+0/19)

7. "public health" "health index" "health monitoring" England OR Wales OR Scotland OR "Northern Ireland" OR “United Kingdom” OR OECD OR Europe OR Canada OR USA OR "United States" OR Austria OR Belgium OR "Czech Republic" OR France OR Germany OR "Great Britain" OR Ireland OR Greece OR Hungary OR Italy OR Netherlands OR Luxemberg OR Poland OR Portugal OR Denmark OR Estonia OR Finland OR Iceland OR Norway OR Sweden OR “Slovak Republic” OR Slovenia OR Spain OR Switzerland OR Turkey OR Israel OR Australia OR “New Zealand” OR Japan OR Korea OR “Organisation for Economic Co-Operation and Development” (n=+8/149)

8. "population health" "health index" "health monitoring" England OR Wales OR Scotland OR "Northern Ireland" OR “United Kingdom” OR OECD OR Europe OR Canada OR USA OR "United States" OR Austria OR Belgium OR "Czech Republic" OR France OR Germany OR "Great Britain" OR Ireland OR Greece OR Hungary OR Italy OR Netherlands OR Luxemberg OR Poland OR Portugal OR Denmark OR Estonia OR Finland OR Iceland OR Norway OR Sweden OR “Slovak Republic” OR Slovenia OR Spain OR Switzerland OR Turkey OR Israel OR Australia OR “New Zealand” OR Japan OR Korea OR “Organisation for Economic Co-Operation and Development” (n=+0/49)

9. "public health" "health indices" "health monitoring" England OR Wales OR Scotland OR "Northern Ireland" OR “United Kingdom” OR OECD OR Europe OR Canada OR USA OR "United States" OR Austria OR Belgium OR "Czech Republic" OR France OR Germany OR "Great Britain" OR Ireland OR Greece OR Hungary OR Italy OR Netherlands OR Luxemberg OR Poland OR Portugal OR Denmark OR Estonia OR Finland OR Iceland OR Norway OR Sweden OR “Slovak Republic” OR Slovenia OR Spain OR Switzerland OR Turkey OR Israel OR Australia OR “New Zealand” OR Japan OR Korea OR “Organisation for Economic Co-Operation and Development” (n=+2/63)

10. "population health" "health indices" "health monitoring" England OR Wales OR Scotland OR "Northern Ireland" OR “United Kingdom” OR OECD OR Europe OR Canada OR USA OR "United States" OR Austria OR Belgium OR "Czech Republic" OR France OR Germany OR "Great Britain" OR Ireland OR Greece OR Hungary OR Italy OR Netherlands OR Luxemberg OR Poland OR Portugal OR Denmark OR Estonia OR Finland OR Iceland OR Norway OR Sweden OR “Slovak Republic” OR Slovenia OR Spain OR Switzerland OR Turkey OR Israel OR Australia OR “New Zealand” OR Japan OR Korea OR “Organisation for Economic Co-Operation and Development” (n=+0/1)

11. "community health" "health indices" "health monitoring" England OR Wales OR Scotland OR "Northern Ireland" OR “United Kingdom” OR OECD OR Europe OR Canada OR USA OR "United States" OR Austria OR Belgium OR "Czech Republic" OR France OR Germany OR "Great Britain" OR Ireland OR Greece OR Hungary OR Italy OR Netherlands OR Luxemberg OR Poland OR Portugal OR Denmark OR Estonia OR Finland OR Iceland OR Norway OR Sweden OR “Slovak Republic” OR Slovenia OR Spain OR Switzerland OR Turkey OR Israel OR Australia OR “New Zealand” OR Japan OR Korea OR “Organisation for Economic Co-Operation and Development” (n=+1/34)

12. "community health" "health index" "health monitoring" England OR Wales OR Scotland OR "Northern Ireland" OR “United Kingdom” OR OECD OR Europe OR Canada OR USA OR "United States" OR Austria OR Belgium OR "Czech Republic" OR France OR Germany OR "Great Britain" OR Ireland OR Greece OR Hungary OR Italy OR Netherlands OR Luxemberg OR Poland OR Portugal OR Denmark OR Estonia OR Finland OR Iceland OR Norway OR Sweden OR “Slovak Republic” OR Slovenia OR Spain OR Switzerland OR Turkey OR Israel OR Australia OR “New Zealand” OR Japan OR Korea OR “Organisation for Economic Co-Operation and Development” (n=0/117)

13. "community health" "health indicators" "health monitoring" England OR Wales OR Scotland OR "Northern Ireland" OR “United Kingdom” OR OECD OR Europe OR Canada OR USA OR "United States" OR Austria OR Belgium OR "Czech Republic" OR France OR Germany OR "Great Britain" OR Ireland OR Greece OR Hungary OR Italy OR Netherlands OR Luxemberg OR Poland OR Portugal OR Denmark OR Estonia OR Finland OR Iceland OR Norway OR Sweden OR “Slovak Republic” OR Slovenia OR Spain OR Switzerland OR Turkey OR Israel OR Australia OR “New Zealand” OR Japan OR Korea OR “Organisation for Economic Co-Operation and Development” (n=+17/1,040)

14. "community health" "mortality indicators" "health monitoring" England OR Wales OR Scotland OR "Northern Ireland" OR “United Kingdom” OR OECD OR Europe OR Canada OR USA OR "United States" OR Austria OR Belgium OR "Czech Republic" OR France OR Germany OR "Great Britain" OR Ireland OR Greece OR Hungary OR Italy OR Netherlands OR Luxemberg OR Poland OR Portugal OR Denmark OR Estonia OR Finland OR Iceland OR Norway OR Sweden OR “Slovak Republic” OR Slovenia OR Spain OR Switzerland OR Turkey OR Israel OR Australia OR “New Zealand” OR Japan OR Korea OR “Organisation for Economic Co-Operation and Development” (n=+2/49)

15. "community health" "morbidity indicators" "health monitoring" England OR Wales OR Scotland OR "Northern Ireland" OR “United Kingdom” OR OECD OR Europe OR Canada OR USA OR "United States" OR Austria OR Belgium OR "Czech Republic" OR France OR Germany OR "Great Britain" OR Ireland OR Greece OR Hungary OR Italy OR Netherlands OR Luxemberg OR Poland OR Portugal OR Denmark OR Estonia OR Finland OR Iceland OR Norway OR Sweden OR “Slovak Republic” OR Slovenia OR Spain OR Switzerland OR Turkey OR Israel OR Australia OR “New Zealand” OR Japan OR Korea OR “Organisation for Economic Co-Operation and Development” (n=+0/24)

| | Ovid Medline | |
|---|---|---|
| **COUNTRY** | **Study name** | **First author (year)** |
| **Multiple** | EURO-HEALTHY ('Shaping EUROpean policies to promote | Costa C (2019), Costa C (2019), Freitas A (2018) |
| | ACT (Advancing Care Coordination and Telehealth programme) | Duenas-Espin I (2016) |
| | EURO-URHIS2 (European Urban Health Indicators System Part | De Gelder R (2017), Koster EM (2010), Pope D (2017) |
| | WHO (World Health Organisation) Healthy Cities | De Leeuw E (2015) |
| | IMOA (Improving Health Monitoring in Old Age) | Grube MM (2019) |
| | EuroMOMO (European Monitoring of Excess Mortality for | Kanieff M (2010) |
| **Australia** | HILDA (Household, Income and Labour. Dynamics in Australia) | Norman R (2013) |
| **Austria** | | |
| **Belgium** | | |
| **Canada** | | |
| **Chile** | | |
| **Colombia** | | |
| **Czech Republic** | | |
| **Denmark** | | |
| **Estonia** | | |
| **Finland** | FINRISK 2007 | Karvanen J (2016) |
| | Multiple | Kilpelainen K (2016) |
| **France** | | |
| **Germany** | DEGS (German Health Interview and Examination Survey for | Scheidt-Nave C (2012) |
| | German Federal Statistical Office | Sundmacher L (2013) |

| | | |
|---|---|---|
| **Greece** | | |
| **Hungary** | | |
| **Iceland** | | |
| **Ireland** | | |
| **Israel** | | |
| **Italy** | Italian Network of Longitudinal Metropolitan Studies (IN- | Caranci N (2018) |
| **Japan** | Japanese Health Statistics Bureau | Rothenberg R (2014) |
| **Korea** | Korean Statistics Bureau | Heo S (2013) |
| | Korean Statistics Bureau | Jung M (2015) |
| **Latvia** | | |
| **Lithuania** | | |
| **Luxembourg** | | |
| **Mexico** | | |
| **Netherlands** | | |
| **New Zealand** | | |
| **Norway** | | |
| **Poland** | | |
| **Portugal** | | |
| **Slovak Republic** | | |
| **Slovenia** | | |
| **Spain** | | |
| **Sweden** | | |
| **Switzerland** | | |
| **Turkey** | | |
| **United Kingdom** | Public Health Metrics | Bellis MA (2012) |
| | Annual Population Survey | Moon G (2019) |
| | 2011 Census | Wheeler BW (2015) |

| | | |
|---|---|---|
| **(England)** | | |
| **(Northern** | | |
| **(Scotland)** | | |
| **(Wales)** | | |
| **United States** | Behavioral Risk Factor Surveillance System (BRFSS) | Asada Y (2014), Dwyer-Lindgren L (2017), Gamble S (2017) |
| | National Health Interview Survey (NHIS) | Jones GC (2013), Zajacova A (2012) |
| | Nurses and Population Health | Fields BE (2016) |
| | Community Health Status Indicators (CHSI) | Kanarek N (2011) |

| Scopus | |
|---|---|
| **Study name** | **First author (year)** |
| EURO-HEALTHY ('Shaping EUROpean policies to promote HEALTH | Santana P (2020) |
| ACT (Advancing Care Coordination and Telehealth programme) | Espieen I (2016) |
| OECD Regional Database | Kim KT (2019) |
| Eurostat Cause of Death database | Weber AC (2017) |
| The Canadian Chronic Disease Surveillance System | Choi B (2013) |
| The Danish Health and Morbidity Survey/Danish National Health Survey | Christensen AI (2020) |
| HRI (German Health Risk Institute) | Andersohn FW (2016) |

| | |
|---|---|
| CSLC (Comprehensive Survey of Living Conditions) | Sugisawa K (2016) |
| Korea Community Health Survey | Kang Y (2015) |
| The Dutch Public Health Monitor | Van de Kassteele J (2017) |
| OCPH 2019 (The Open Comparisons Public Health Study) | Makenzius M (2019) |

| | |
|---|---|
| The Scottish Health and Ethnicity Linkage Study | Bhopal RF (2011) |
| AHRs (America's Health Rankings) | Erwin P (2011) |
| Medical Expenditure Panel Survey Data | Jerant AF (2012) |
| The Rankings (County Health Rankings & Roadmaps) | Park HR (2015) |
| CDC Wonder Programme | Shandera W (2014) |

| Google Scholar | |
|---|---|
| **Study name** | **First author (year)** |
| Czech Statistical office | Hubelova D (2018) |
| Health 2011 | Aromaa A (2019) |
| GEDA (German Health Update) | Lange C (2015), Lange C (2017), Saß AC (2017) |

| | |
|---|---|
| 2nd Dutch National Survey of General Practice | Kroneman M (2010) |
| Inquerito Nacional de Saude com Exame Fisico | Nunes B (2019) |

| | |
|---|---|
| BRFSS | Nguyen T (2017) |
| 500 Cities project | Gourevitch MN (2019) |

| Heallth Interview Survey | |
|---|---|
| **Study name** | **URL** |
| European Health Interview Study (EHIS) | https://ec.europa.eu/eurostat/web/microdata/european-health-interview-survey |
| Australian Health Survey | https://www.abs.gov.au/australianhealthsurvey |
| EHIS | |
| EHIS /Belgian Health Interview Survey | https://hisia.wiv-isp.be/SitePages/Home.aspx |
| Canadian Community Health Survey | https://www.statcan.gc.ca/eng/survey/household/3226 |
| Encuesta Nacional de Salud (ENS) | http://ghdx.healthdata.org/record/chile-national-health-survey-2016-2017 |
| Encuesta Nacional De Salud | http://epi.minsal.cl/wp-content/uploads/2018/05/ENS_F1_corr8Mayo.pdf |
| EHIS | |
| EHIS | |
| EHIS | http://pxweb.tai.ee/PXWeb2015/Resources/PX/Databases/05Uuringud/01ETeU/01Ter |
| EHIS | |
| EHIS | |
| EHIS/DEGS/GEDA | https://www.rki.de/EN/Content/Health_Monitoring/HealthSurveys/HealthSurveys_nod |

| | |
|---|---|
| EHIS | |
| EHIS | |
| EHIS | |
| EHIS | |
| The Israeli National Health Interview Survey | https://www.health.gov.il/English/MinistryUnits/ICDC/Health_Surveys/Pages/INHIS.aspx |
| EHIS | |
| Korea Community Health Survey | https://www.cdc.go.kr/cdc_eng/ |
| EHIS | |
| EHIS | |
| EHIS | |
| EHIS | |
| New Zealand Health Survey | https://www.health.govt.nz/nz-health-statistics/national-collections-and-surveys/surveys/new-zealand-health-survey |
| EHIS | |
| EHIS | |
| EHIS | |
| EHIS | |
| EHIS | |
| EHIS/ European Health Interview for Spain | https://www.ine.es/en/metodologia/t15/t153042014_en.pdf |
| EHIS | |
| Swiss Health Survey | https://www.bfs.admin.ch/bfs/en/home/statistics/health/state-health.html |
| Demographic and Health Survey | https://dhsprogram.com/methodology/survey/survey-display-548.cfm |

| | |
|---|---|
| Health Survey for England | https://digital.nhs.uk/data-and-information/publications/statistical/health-survey-for-england |
| The Health Survey Northern Ireland | https://www.health-ni.gov.uk/topics/doh-statistics-and-research/health-survey-northern- |
| Scottish Health Survey | https://www2.gov.scot/Topics/Statistics/Browse/Health/scottish-health-survey |
| The Welsh Health Survey/National Survey for Wa | https://gov.wales/national-survey-wales |
| National Health Interview Survey (NHIS) | https://www.cdc.gov/nchs/nhis/index.htm |

| Heallth Examination Survey | |
|---|---|
| **Study name** | **URL** |
| Encuesta Nacional de Salud (ENS) | http://ghdx.healthdata.org/record/chile-national-health-s |
| Encuesta Nacional de Situación Nutricional (ENSIN) | http://ghdx.healthdata.org/record/colombia-demographic-and-health-survey-2015-2016 |

| | |
|---|---|
| The Japan National Health and Nutrition Survey | https://www.nibiohn.go.jp/eiken/english/research/projec |
| The Korea National Health and Nutrition Examination | https://knhanes.cdc.go.kr/knhanes/eng/index.do |
| Mexico Health and Nutrition Survey (ENSANUT) | https://ensanut.insp.mx/ |
| Inquerito Nacional de Saude come Exame Fisico | http://www.insef.pt/English/Pages/Inicio.aspx |

| | |
|---|---|
| Health Survey for England | https://digital.nhs.uk/data-and-information/publications/statistical/health-survey-for-england |

| Statistical Agency | |
|---|---|
| NAME | URL |
| Eurostat | |
| Australian Bureau of Statistics | https://www.abs.gov.au/ |
| Statistics Austria | https://www.statistik.at/web_en/statistics/index.html |
| StatBel | https://statbel.fgov.be/en |
| Canadian Vital Statistics: Death Database (CVSD) | https://www.statcan.gc.ca/eng/start |
| Insituto Nacional de Estadisticas | https://www.ine.es/en/index.htm |
| National Administrative Department of Statistics (DANE) | http://www.ins.gov.co/Paginas/Inicio.aspx |
| Czech Statistical Office | https://www.czso.cz/csu/czso/home |
| Statistics Denmark | https://www.dst.dk/en |
| Statistics Estonia | https://www.stat.ee/en |
| StatFin | https://www.stat.fi/tup/statfin/index_en.html |
| Institut national de la statistique et des etudes economiques | https://www.insee.fr/langue/en |
| DeStatis | https://www.destatis.de/EN/Home/_node.html |

| | |
|---|---|
| Hellenic Statistical Agency | https://www.statistics.gr/en/home/ |
| Hungarian Central Statistical Office | http://www.ksh.hu/?lang=en |
| Statistics Iceland | https://www.statice.is/ |
| Central Statisics Office | https://www.cso.ie/en/index.html |
| Central Bureau of Statistics | https://www.cbs.gov.il/en/Pages/default.aspx |
| Instituto Nazionale di Staistics (Istat) | https://www.istat.it/en/ |
| Statistics Bureau of Japan | https://www.stat.go.jp/english/ |
| Korean Statistical Information Service (KOSIS) | https://kosis.kr/eng/ |
| Central Statistical Bureau of Latvia | https://www.csb.gov.lv/en/ |
| Staistics Lithuania | https://osp.stat.gov.lt/statistiniu-rodikliu-analize?region=all# |
| STATEC | https://statistiques.public.lu/en/index.html |
| National Institute of Statistics and Geography (INEGI) | https://en.www.inegi.org.mx/ |
| Statistics Netherlands (CBS) | https://www.cbs.nl/en-gb |
| Stats NZ | https://www.stats.govt.nz/ |
| Norwegian Institute of Public Health | https://www.fhi.no/en/ |
| Statistics Poland | https://stat.gov.pl/en/ |
| Instituto Nacional De Estatistica | https://www.ine.pt/xportal/xmain?xpgid=ine_main&xpid=I |
| Statistical Office of the Slovak Republic | http://datacube.statistics.sk/#!/view/en/VBD_SK_WIN/zd3 |
| Statistical Office of the Republic of Slovenia (SiStat) | https://www.stat.si/Statweb/En |
| Instituto Nacional de Estadistica (INE) | https://www.ine.es/en/ |
| Statistics Sweden National Board of Health and Wefare | https://www.socialstyrelsen.se/en/ |
| The Swiss Federal Statistical Office | https://www.bfs.admin.ch/bfs/en/home.html |
| Turkish Statistical Institute (TurkSTAT) | https://www.tuik.gov.tr/Home/Index# |
| Office for National Statistics (ONS) | www.ons.gov.uk |

| | |
|---|---|
| Office for National Statistics (ONS) | www.ons.gov.uk |
| Northern Ireland Statistics and Research Agency (NIRA) | https://www.health-ni.gov.uk/ |
| National Records of Scotland | https://www.scotpho.org.uk/ |
| statsWales | https://statswales.gov.wales/Catalogue |

| Public health institute | |
|---|---|
| **NAME** | **URL** |
| Australian Institute of Health and Welfare | https://www.aihw.gov.au/ |

| |
|---|
| / |

| | |
|---|---|
| Department of Health | https://www.health-ni.gov.uk/ |
| The Scottish Public Health Observatory | https://www.scotpho.org.uk/ |
| Centers for Disease Control (CDC) | https://wonder.cdc.gov/ |

| Other webite | |
|---|---|
| **NAME** | **URL** |
| European Health & Life Expectancy Information System | http://www.eurohex.eu/index.php?option=ehleisproject |
| National Mortality Database | https://www.aihw.gov.au/about-our-data/our-data-collections/national-mortality-database |
| Finnish cause of death register | https://www.stat.fi/meta/til/ksyyt_en.html |
| Robert Koch Institute | https://www.rki.de/EN/Home/homepage_node.html |

| | |
|---|---|
| American Community Survey (ACS) | https://www.census.gov/programs-surveys/acs |

**Supplemental Table 5. Health indicators available in OECD countries [a] by sub-country geographies: morbidity**

| Country | Health indicator data source | Years data available | Age range | Self-rated health | Long-standing illness | Disabled | Activity limitation | Healthy Life Exp. | Geographic data boundary available |
|---|---|---|---|---|---|---|---|---|---|
| Europe (28 countries) | EURO-HEALTHY [b] | 2000-15 | Varies | √ | √ | √ (DALYs) | | | Region (NUTS 2) |
| | EU-SILC | 2003-2018 | 16+ | √ | √ | | √ | | Region (NUTS 1) |
| | EHIS | 2006-9, 2013-15 | 15+ | √ | √ | | √ | √ | Region (NUTS 2) |
| Europe (14 countries) | EURO-URHIS2 [c] | 2010-2011 | Varies | √ | | | | | Urban area |
| Australia | HILDA | 2009-2010 | 18+ | | | | | | Areas |
| | Australian Health Survey | 2011-2013 | All | √ | √ | √ | | | State/territory |
| | Census | 2006, 2011, 2016 | All | | | √ | | | Many (e.g. SA1s) |
| Canada | CCHS | 2001-2020 | 12+ | √ | | | √ | | Province or territory/ metropolitan area |
| | CCDSS | 2004-2006 | All | | | | | √ | Province or territory |
| | Canadian Survey on Disability | 2017 | 15+ | | | √ | | | Province or territory/ metropolitan area |
| Chile | Encuesta Nacional de Salud (ENS) | 2003, 2009-10, 2016-17 | 15+ | √ | | | | | Region |
| Colombia | ENSIN | 2005, 2010, 2015 | 0-64 | √ | | | | | Region |
| | SABE | 2015 | 60+ | √ | √ | | √ | | Region |
| Denmark | Danish National Health Survey | 1987, 1994, 2000, 2005, 2010, 2013, 2017 | 16+ | √ | √ | | | | Region |
| Estonia | Statistics Estonia/ Census | 2011 | All | | √ | | √ | √ | Region |
| Finland | The Regional Health and Wellbeing Study | 2012-2014, 2017 | 20+ | √ | | | | | Region/ Municipality (>20,000) |
| | Health 2011 | 2011 | 25+ | √ | √ | | | | Region [d] |
| | FINRISK/ FinHealth | 2012/ 2017 | 25+ | √ | √ | | | | Region |
| Germany | German Health Update (GEDA) | 2009, 2010, 2012, 2014/5 | 15+ | √ | √ | √ | √ | | Region/ Municipality |
| | DEGS | 2008-2011 | 18+ | √ | √ | √ | | | Region |
| | SOEP | 1984-2020 | All | √ | | | | | Region |
| Hungary | Hungarian CSO/ Census | 2011 | All | | √ | | | | Region/ County |

| | | | | | | | | | |
|---|---|---|---|---|---|---|---|---|---|
| Ireland | CSO/ Census | 2011, 2016 | All | √ | | √ | | | Region/County/City |
| Italy | Aspects of Daily Life survey | 1993-2003, 2005-18 | All | √ | √ | | | | Municipality |
| Japan | Statistics of Japan, CSLC | 2010-2018 | All | √ | √ | | | | District/ Prefecture |
| Lithuania | L-SILC | 2005-2019 | 18-64 | | | √ | | | Region/ County |
| Mexico | ENSANUT | 2006, 2012, 2016, 2018 | All | √ | √ | | | | Administrative territory |
| The Netherlands | ECHIM | 2011-2013 | Varies | √ | √ | | √ | √ | Region |
| | Dutch Public Hlth Monitor | 2012 | 19+ | √ | √ | √ | | | Municipality/District |
| | 2nd Dutch NSGP | 2000-2002 | All | √ | √ | | | | 4-digit postcode |
| New Zealand | New Zealand Health Survey | 2002/3, 2006/7, 2011-2020 | All | √ | | | | | Region/district/public health unit/PSUs |
| | Census | 2018 | All | | | | √ | | Region |
| Portugal | INSEF | 2015 | 25-74 | √ | √ | | √ | | Region/Urban area |
| Slovak Republic | Statistical Office of the Slovak Republic | 2001-2018 | 'Working age' | | | √ (to work) | | | Region (nuts 2) |
| Spain | INE | 2008 | 6+ | | | √ | | √ [e] | AC/ Municipality |
| South Korea | KCHS | 2008-2020 | 19+ | √ | | | √ | | City/county/ District |
| Sweden | OCPH | 2019 | 16-84 | √ | | | | | Region/Municipality |
| USA | ACS | 2013-2019 | All | | | √ | | | Tract/block group [f] |
| | BRFSS | 2002-2017 | 18+ | √ | | | √ | | State/Metropolitan/ Micropolitan/County |
| | The Rankings | 2010-2013 | All | √ | | | | | County |
| | CHSI | 2008 | All | √ | | | | | County (>100,000) |
| | NHIS | 1986-2020 | All | √ | | √ | | | block, block group, tract, county, state |
| United Kingdom | Census [g] | 2011 | All | √ | √ | | | | Local Authority / OA/Data Zone |
| | ONS | 2009/11, 2016/18 | All | | | √ | | √ | Local Authority |
| (England) | HSE | 2004-2018 | All | √ | √ | | √ | | Region |
| (Northern Ireland) | Department of Health | 2012/14, 2016/18 | All | | | | | √ | Region/Trust/District |
| (Scotland) | Scottish Health Survey (SHS) | 1995, 1998, 2003, 2008-2018 | All | √ | √ | | | | Health board/ Local Authority (2008+) |
| | The Scottish Public Health Observatory | 2015/16, 2016/18 | All | | | | | √ | Region/ NHS Board/ Local Authority [h] |
| (Wales) | WHS/NSW | 2003-2013, 2016-2020 | 16+ | √ | √ | | | | Health board/ Local Authority |

Abbreviations: SILC, Survey on Income and Living Conditions; EHIS, European Health Interview Survey; EURO-URHIS2, European Urban Health Indicators Part Two; HILDA, Household, Income and Labour Dynamics in Australia; CCHS, Canadian Community Health Survey; CCDSS, Canadian Chronic Disease

Surveillance System; ENSIN, Encuesta Nacional de Situación Nutricional; SABE, Survey on Health, Well-being and SAlud, Bienestar & Envejecimiento; DEGS, German Health Interview and Examination Survey for Adults; SOEP, German Socio-Economic Panel; CSO, Central Statistical Office; CSLC, Comprehensive Survey of Living Conditions; ENSANUT, Mexico Health and Nutrition Survey; NSGP, National Survey of General Practitioners; INSEF, Inquerito Nacional de Saude come Exame Fisico; INE, Instituto Nacional de Estadistica; KCHS, Korean Community Health Survey; OCPH, The Open Comparisons Public Health study; ACS, American Community Survey; BRFSS, Behavioral Risk Factor Surveillance System; CHSI, Community Health Status Indicators; NHIS, National Health Interview Survey; HSE, Health Survey for England; WHS, The Welsh Health Survey; NSW, National Survey for Wales.]

[a] No overall health indicator data in English below country-level for Israel and Turkey.
[b] Austria, Belgium, Denmark, Estonia, France, Germany, Greece, Hungary, Ireland, Italy, Luxembourg, Malta, Netherlands, Norway, Portugal, Spain, Sweden, Switzerland and United Kingdom.
[c] France, Germany, The Netherlands, United Kingdom, Slovenia, Norway, Slovakia, Romania, Former Macedonia, Turkey, Latvia and Lithuania.
[d] 20 strata, made up of 15 of largest cities and rest country divided into 5 regions.
[e] Only at Autonomous Community level in 2008.
[f] Geographies of 20,000 people, or more, also available 2008-2012.
g Lower Super Output Areas (LSOA) and Middle Super Output Areas (MSOA) in England/Wales; Data zones for Scotland.
[h] Plus a ‘Burden of disease’ measure.

| | | | | | | | |
|---|---|---|---|---|---|---|---|
| | | | MORTALITY | | | | |
| **COUNTRY *** | **All-cause** | **Cause-specific** | **Life Exp. birth** | **Life Exp. 65y** | **Preventable** | **Excess** | **Amenable** |
| Australia | R/ PHU/ SA3 | | | | R/ PHU/ SA3 | | |
| Austria | R2/ R3/ P/ UA | UA | R2 | | R2 | R2 | R2 |
| Belgium | R2/ UA/ P/ Di | R2/ Mu/ UA | R2/ Mu | | R2/ Mu | R2 | R2/Mu |
| Canada | P/ T/ PHR/ PHU | P/ T/ PHR/ PHU | P/T/PHR/PHU | P/T/PHR/PHU | P/T/PHR/PHU | | |
| Chile | | | | | | | |
| Colombia | | | | | | | |
| Czech Republic | R2/ Mu/ UA/ Di | R2/ Mu/ UA/ Di | R2/ Mu | | R2/ Mu | R2 | R2/Mu |
| Denmark | R2/ Mu/ UA/ P | R2/ Mu/ UA/ P | R2 | | R2 | R2 | R2 |
| Estonia | R2/ UA/ Co | R2/ UA | R2/ Co | | R2 | R2 | R2 |
| Finland | R2/ Mu/ UA | R2/ Mu/ UA | R2 | | R2 | R2 | R2 |
| France | R1/ R2/ R3/ Mu/ UA | R1/ R2/ R3/ Mu/ UA | R1/R2/R3/Mu | R1/ R2/ R3 | R2/ Mu | R2 | R2/Mu |
| Germany | R1/R2/ Mu/ UA/ Di | R1/R2/ Mu/ UA/ Di | R2/ Mu/ Di | | R2/ Mu/ Lander/ Di | R2 | R2/Mu |
| Greece | R2/ R3/ Mu/ UA/ Pr | R2/ R3/ Mu/ UA/ Pr | R2/ R3/ Mu | | R2/ Mu | R2 | R2/Mu |
| Hungary | R2/ UA/ Co | R2/ UA/ Co | R2 | | R2 | | R2 |
| Iceland | R2/ UA/ Mu | R2/ UA | R2 | | R2 | R2 | R2 |
| Ireland | R2/ UA | R2/ UA | R2 | | R2 | | R2 |
| Israel | Di/ SubDi | | | | | R2 | |
| Italy | R2/ Mu/ UA/ Pr | R2/ Mu/ UA/ Pr | R2/ Mu | R / P/ Mu | R2/ Mu | | R2/Mu |
| Japan | Ci/ Pr | Ci/ Pr | Ci/ Pr | Ci/ Pr | | | |
| Korea | P/ Ci/ Di | P/ Ci/ Di | | | | | |
| Latvia | R2/ UA/ Co/ Ci | R2/ UA | R2/ Co/ Ci | | R2 | | R2 |
| Lithuania | R2/ UA/ Co | R2/ UA/ Co | R2 | | R2 | R2 | R2 |
| Luxembourg | R2 | R2 | R2 | | R2 | | R2 |
| Mexico | St | St | St | | | St | |
| Netherlands | R2/ UA | R2/ UA | R2 | | R2 | | R2 |
| New Zealand | R/ Di/ HB/ LB | | R/ Di/ HB | | | R2 | |

| | | | | | | | |
|---|---|---|---|---|---|---|---|
| Norway | R2/ UA/ Co/ Ci | R2/ UA | R2/ Co/ Ci | | R2 | | R2 |
| Poland | R2/ UA | R2/ UA | R2 | | R2 | R2 | R2 |
| Portugal | R2/ R3/ Mu | R2/ Mu/ UA | R2/ R3/ Mu | R3 | R2/ Mu | | R2/Mu |
| Slovak Republic | R2/ R3/ Mu/ UA/ Di | R2/ R3/ Mu/ UA/ Di | R2 | | R2 | | R2 |
| Slovenia | R2/ Mu/ UA | R2/ UA | R2/ Mu | | R2 | R2 | R2 |
| Spain | R2/ Mu/ UA/ AC | R2/ Mu/ UA/ AC | R2/ Mu/ AC | | R2/ Mu | R2 | R2/Mu |
| Sweden | R2/ Mu/ UA/ Co | R2/ Mu/ UA/ Co | R2/ Mu/ Co | | R2/ Mu | R2 | R2/Mu |
| Switzerland | R2/Di/ Canton/Com | R2 | R2 | | R2 | | R2 |
| Turkey | R/ subR/ P/ UA | R/ subR/ P/ UA | | | | R2 | |
| United Kingdom | R2/UA/Mu/LA/CCG/HB | R2/UA/Mu/LA/CCG/HB | R2/Mu/LA/CCG/ HB | R2/LA/CCG/HB | R2/Mu/LA/CCG/HB | R2/Mu/LA/CCG/HB | R2/Mu |
| United States | St/ Co/ Ci | St/ Co/ Ci | | | | R2 | |

Abbreviations: AU, Autonomous Community; CCG, Clinical Commissioning Group; Co, County; Com, Commune; Di, District; DZ, Data Zone; HB, H[illegible] Authority; LB, Local Board; Me, Metropolitan Area; Mu, Municipality; P, Province; PHR, Public Health Region, PHU, Public Health Unit; Pr, Prefectu[illegible] NUTS or unspecified; R1, Region NUTS 1; R2, Region NUTS2; R3, NUTS 3; OA, Output Area; St, State; T, Territory; UA, Urban Area.

* All OECD countries have data on all-cause mortality and Life Expectancy at birth at geographic levels TL2 and TL3, and some causes of death data at

| MORBIDITY | | | | |
| --- | --- | --- | --- | --- |
| **Self-rated health** | **Long-standing illness** | **Disabled** | **Activity limitation** | **Healthy Life Expectancy** |
| St/ T | St/ T | St/ T/ Other | | |
| R1/ R2/ UA/ Mu | R1/ R2/ UA/ Mu | R2 | R1/ R2/ UA | R2 |
| R1/ R2/ UA/ Mu | R1/ R2/ UA/ Mu | R2/ Mu | | R2 |
| Province/ T/ Me | | Province/ T/ Me | Province/ T/ Me | Province/ T |
| R | | | | |
| R | R | | R | |
| R1/ R2/ UA/ Mu | R1/ R2/ UA/ Mu | R2/ Mu | R1/ R2/ UA/ Mu | R2 |
| R1/ R2/ UA | R1/ R2 | R1/ R2 | R1/ R2/ UA | R2 |
| R1/ R2/ UA | R1/ R2/ UA | R2 | R1/ R2/ UA | R2 |
| R1/ R2/ UA | R1/ R2/ UA | R2 | R1/ R2/ UA | R2 |
| R1/ R2/ UA/ Mu | R1/ R2/ UA/ Mu | R2/ Mu | R1/ R2/ UA/ Mu | R2 |
| R2/ UA/ Mu | R2/ UA/ Mu | R2/ Mu | R2/ UA/ Mu | R2 |
| R1/ R2/ UA/ Mu | R1/ R2/ UA/ Mu | R2/ Mu | R1/ R2/ UA/ Mu | R2 |
| R1/ R2/ UA | R1/ R2/ UA/Co | R2 | R1/ R2/ UA | R2 |
| R1/ R2/ UA | R1/ R2/ UA | R2 | R1/ R2/ UA | R2 |
| R1/ R2/ UA/Co/Ci | R1/ R2/ UA | R2/ Co/ City | R1/ R2/ UA | R2 |
| R1/ R2/ UA/ Mu | R1/ R2/ UA/ Mu | R2/ Mu | R1/ R2/ UA/ Mu | R2 |
| Di | Prefecture | | | |
| Co/ Ci/ Di | | | Co/ Ci/ Di | |
| R1/ R2/ UA | R1/ R2/ UA | R2 | R1/ R2/ UA | R2 |
| R1/ R2/ UA | R1/ R2/ UA | R2/ Co | R1/ R2/ UA | R2 |
| R1/ R2 | R1/ R2 | R2 | R1/ R2 | R2 |
| Administrative T | Administrative T | | | |
| R1/ R2/ UA/ Mu/ Di | R1/ R2/ UA/ Mu/ Di | R2/ Mu/ Di | R1/ R2/ UA | R2 |
| R/ Di/ PHU | | | R | |

| | | | | |
|---|---|---|---|---|
| R1/ R2/ UA | R1/ R2/ UA | R2 | R1/ R2/ UA | |
| R1/ R2/ UA | R1/ R2/ UA | R2 | R1/ R2/ UA | R2 |
| R1/ R2/ Mu/ UA | R1/ R2/ Mu/ UA | R2/ Mu | R1/ R2/ UA | R2 |
| R1/ R2/ UA | R1/ R2/ UA | R2 | R1/ R2/ UA | R2 |
| R1/ R2/ UA | R1/ R2/ UA | R2 | R1/ R2/ UA | R2 |
| R1/ R2/ UA/ Mu | R1/ R2/ UA/ Mu | R2/ Mu/ AC | R1/ R2/ UA/ Mu | R2/ Mu/ AC |
| R1/ R2/ UA/ Mu | R1/ R2/ UA/ Mu | R2/ Mu | R1/ R2/ UA/ Mu | R2 |
| R1/ R2 | R1/ R2 | R2 | R1/ R2 | |
| R1/ R2 | R1/ R2 | | R1/ R2 | |
| R1/ R2/ UA/ Mu/ LA/OA-DZ | R1/ R2/ UA/ Mu/ LA/OA-DZ | R2/ Mu/ LA | R1/ R2/ UA/ Mu | R2/ Mu/ LA |
| S/ Co/ Me/ Mi/ Tract/ BG/ B | | St/ Co/ Tract/ BG/ B | St/ Me/ Mi/ Co | |

ealth Board; LA, Local
re; R, Region not

geographic level TL2
